# Cerebrospinal fluid α-synuclein seed amplification assay positivity identifies a prognostically distinct subgroup in amyotrophic lateral sclerosis

**DOI:** 10.64898/2026.10.23.26366349

**Authors:** Kazuki Obara, Andrea Mastrangelo, Agustina Ranieri, Shorena Janelidze, Daisuke Ito, Munetaka Yamamoto, Veria Vacchiano, Rocco Liguori, Serena Caldera, Yuichi Riku, Yasushi Iwasaki, Sinh Tran, Takahiro Kawase, Yu Mori, Keita Hiraga, Shinichiro Yamada, Koyo Tsujikawa, Yosuke Nishio, Tomoo Ogi, Ryoichi Nakamura, Naoki Atsuta, Gen Sobue, Erik Stomrud, Sebastian Palmqvist, Oskar Hansson, Piero Parchi, Masahisa Katsuno, Niklas Mattsson-Carlgren

**Author notes:** These authors jointly supervised this work (Masahisa Katsuno, Niklas Mattsson-Carlgren). **Correspondence to**: **Kazuki Obara**, Clinical Memory Research Unit, Department of Clinical Sciences Malmö, Lund University. BMC C11, Baravägen 18, 22241 Lund, Sweden., **Correspondence may also be addressed to: Masahisa Katsuno**, Department of Neurology, Nagoya University Graduate School of Medicine 65 Tsurumai-cho, Showa-ku, Nagoya, 466-8550, Japan., **Correspondence may also be addressed to: Niklas Mattsson-Carlgren**, Memory Clinic, Skåne University Hospital, Malmö 20122, Sweden.

## Abstract

**Background:** α-synuclein (Lewy body-type) co-pathology has been identified at autopsy in a subset of patients with amyotrophic lateral sclerosis (ALS). However, its prevalence and prognostic relevance *in vivo* remain poorly characterized.

**Methods:** We analyzed baseline cerebrospinal fluid (CSF) α-synuclein seed amplification assay (αSyn SAA) status in 443 patients with ALS from two cohorts (Nagoya, Japan, *n* = 248; Bologna, Italy, *n* = 195). Samples were classified as strong-positive, weak-positive, or negative based on RT-QuIC reactivity across replicate wells, using the assay’s established *a priori* criteria at initial and repeat testing; classifications were corroborated by quantitative kinetic parameters. We compared age– and sex-adjusted odds of positivity in ALS with those in cognitively unimpaired (CU; *n* = 999; stratified by amyloid-β [Aβ] status), Aβ-positive mild cognitive impairment (*n* = 224), and Alzheimer’s disease dementia (*n* = 319) groups from the Swedish BioFINDER cohort. In ALS, we examined tracheostomy-free survival and longitudinal functional decline using cohort-stratified Cox and linear mixed-effects models, respectively. We also assessed interactions between αSyn SAA status and baseline CSF neurofilament light chain (NfL). Prognostic analyses were performed in sporadic ALS and repeated after including familial or genetic cases.

**Results:** Strong αSyn SAA positivity was detected in 24/443 patients with ALS (5.4%); overall positivity, including weak-positive samples, was 37/443 (8.4%). Patients with ALS did not have significantly higher odds of SAA positivity than Aβ-negative CU participants (age– and sex-adjusted odds ratio [OR] 1.33, 95% CI [0.82, 2.14]) and had significantly lower odds than patients with Alzheimer’s disease dementia (OR 0.44, [0.27, 0.70]). Among patients with sporadic ALS, strong αSyn SAA positivity was associated with shorter tracheostomy-free survival after adjustment for established predictors including NfL (adjusted HR 2.08 [1.25, 3.45]), whereas weak positivity showed no clear survival association. In linear mixed-effects models, strong αSyn SAA positivity modified the association between NfL and ALSFRS-R decline (NfL × αSyn SAA × time interaction β−0.70 points/month [−1.26, −0.14] per SD NfL). These findings persisted in analyses including familial or genetic ALS cases and in those restricted to Aβ-negative patients. Neuropathological findings (*n* = 5) were concordant with the CSF αSyn SAA classification.

**Conclusions:** A strong α-synuclein seeding profile was uncommon in ALS but identified a subgroup with shorter survival and a stronger association between NfL and functional decline. These findings provide ante-mortem evidence that α-synuclein co-pathology may contribute to prognostic heterogeneity in ALS.

## Introduction

Amyotrophic lateral sclerosis (ALS) is a progressive and fatal neurodegenerative disorder, characterized by a highly heterogeneous clinical course.^1,2^ This heterogeneity complicates prognostic counselling and the design of clinical trials. Neurofilament light chain (NfL) is well validated as a prognostic biomarker across ALS cohorts; however, it is not disease-specific and does not fully capture the clinical trajectory of ALS.^3^

One potential contributor to heterogeneity is the presence of co-existing neurodegenerative pathologies. While transactive response DNA binding protein 43 kDa (TDP-43)-proteinopathy is the defining neuropathological feature of sporadic ALS,^2,4^ additional proteinopathies including amyloid-β (Aβ) are increasingly recognized,^5–7^ and may contribute to biological and clinical heterogeneity.^8,9^ We previously reported that CSF Aβ positivity status modified the relationship between NfL and the rate of functional decline in ALS, particularly among participants with high NfL values.^10^ These findings raised the broader possibility that co-existing neurodegenerative pathologies may define prognostically distinct ALS subgroups.

α-synuclein (Lewy body-type) co-pathology has been described at autopsy in subsets of ALS patients,^5,6^ but whether it represents an incidental age-related finding or contributes to clinical heterogeneity and disease progression remains unclear. CSF α-synuclein seed amplification assays (αSyn SAAs) now enable specific ante-mortem detection of misfolded α-synuclein seeds and have transformed the biomarker landscape of synucleinopathies.^11,12^ Beyond binary positivity, the degree and consistency of SAA reactivity may provide additional information on α-synuclein seed burden, with stronger seeding signals reported in more widespread Lewy body pathology.^13,14^ Their application has since expanded across the neurodegenerative disease spectrum, particularly in Alzheimer’s disease (AD).^15^ More recently, misfolded α-synuclein seeds were also detected in post-mortem CSF from a subset of patients with ALS.^9^ However, the prevalence and prognostic significance of *in vivo* αSyn SAA positivity in ALS remain largely uncharacterized. Importantly, survival associations derived from post-mortem studies are difficult to interpret because patients with slower disease progression are more likely to survive long enough to harbour incidental age-related co-pathologies at death. Ante-mortem CSF αSyn SAA study can address these gaps by establishing α-synuclein status before subsequent clinical outcomes are observed.

We therefore aimed to assess (i) the prevalence of *in vivo* CSF αSyn SAA positivity in ALS relative to controls, (ii) its associations with survival and longitudinal functional decline, and (iii) whether αSyn SAA status modifies the association between CSF NfL and prognosis.

## Materials and methods

### Participants

#### ALS cohorts

The study population was derived from two independent ALS cohorts: the Bologna cohort (Bologna, Italy; enrolled between September 2014 and July 2025) and the Nagoya cohort (Nagoya, Japan; enrolled between January 2013 and September 2024).^8,10^ At each participating site, a diagnosis of ALS was established by neurologists and neurophysiology specialists in accordance with the revised El Escorial criteria.^16^ Participants from both cohorts provided a baseline CSF sample and were followed prospectively to ascertain survival and assess functional decline using the revised Amyotrophic Lateral Sclerosis Functional Rating Scale (ALSFRS-R).^17^ We included consecutive patients with ALS who had a baseline CSF sample with a valid αSyn SAA result from both cohorts.

#### BioFINDER cohort

Comparator participants were drawn from the prospective Swedish BioFINDER-2 study (NCT03174938), which enrols participants from the general population as well as from the memory and neurology clinics of Skåne University Hospital and Ängelholm Hospital in southern Sweden; its full eligibility criteria have been reported previously.^18^ Cognitive classification followed the procedures established for this cohort.^18^ Briefly, participants presenting with cognitive symptoms completed extensive neuropsychological batteries, and were designated as having mild cognitive impairment (MCI) when performance in at least one domain fell ≥1.5 SD below age– and education-adjusted norms; those performing above this threshold were designated as having subjective cognitive decline (SCD). Participants with intact cognition and those with SCD were analysed together as a single cognitively unimpaired (CU) group, none of whom fulfilled the clinical criteria for Parkinson’s disease or prodromal dementia with Lewy bodies at baseline.^19,20^ A diagnosis of AD dementia required both CSF Aβ positivity and fulfilment of DSM-5 criteria for dementia due to AD. Aβ status was determined from the CSF Aβ42/Aβ40 ratio, as described below. For the present analysis we retained CU, Aβ-positive MCI, and Aβ-positive AD dementia participants with baseline CSF αSyn SAA result and an Aβ42/Aβ40 measurement.

### CSF biomarker measurement

CSF Aβ42/40 ratio was quantified using Lumipulse G assays (Fujirebio) in the Bologna cohort and the V-PLEX Aβ Peptide Panel 1 (6E10) Kit on the MSD platform (Meso Scale Diagnostics) in the Nagoya cohort. Aβ positivity was defined by pre-specified, cohort-specific cut-offs (ratio < 0.068 for Bologna; ratio < 0.077 for Nagoya).^8,10^ In BioFINDER cohort, CSF Aβ42 and Aβ40 were measured using the Elecsys CSF electrochemiluminescence immunoassay (Roche Diagnostics) in most participants (77.6%, *n* = 1,197) and Lumipulse G (Fujirebio; *n* = 331) or MSD (*n* = 14) assays in the others. Aβ positivity was defined by previously described assay-specific cut-offs for the Aβ42/40 ratio (< 0.080 for Elecsys; < 0.072 for Lumipulse; < 0.077 for MSD).^21,22^

In the Bologna cohort, CSF NfL values were obtained using the Simoa SR-X platform (NfL Advantage Assay, Quanterix; *n* = 153) or the Lumipulse platform (*n* = 42). Lumipulse values were bridged to Simoa-equivalent values using linear regression of log-transformed measurements from all available samples assessed on both platforms (*n* = 48; Pearson *r* = 0.97). No extrapolation beyond the paired-sample range was required. In the Nagoya cohort, all CSF NfL measurements were performed using the Simoa SR-X platform (Neurology 2-Plex B Assay, Quanterix).^10^

#### CSF αSyn SAA (RT-QuIC)

All CSF αSyn SAA values were analyzed using an established Real-Time Quaking-Induced Conversion (RT-QuIC) protocol with four replicates per sample, at the same laboratory at the University of Bologna.^23^ Initial runs yielding ≥3 positive wells out of 4 were classified as *strong-positive*, representing a high seed burden.^21^ Samples with 1–2 reactive wells underwent two additional runs (12 replicates total), with ≥4 reactive wells across all runs required for classification as *weak-positive*; all other samples were categorized as *negative*.^14^ Kinetic parameters, including maximum fluorescence intensity (Imax) and lag-phase duration (lag time),^24^ were extracted according to established protocols^24^ and standardized within cohorts to minimize batch-related variability.

### Clinical assessments and outcomes

The primary prognostic outcome was defined as the duration from baseline assessment to either death or tracheostomy, with survivors censored at the date of last contact. For longitudinal assessment, functional status was also tracked using serial total ALSFRS-R scores. Baseline clinical and demographic variables included the site of onset (categorized as bulbar/respiratory or spinal), disease duration (months), the baseline progression rate (ΔFS; determined as [48 − baseline ALSFRS-R] / disease duration), forced vital capacity (FVC, percent predicted), and the results of genetic screening for ALS-associated mutations (e.g., *SOD1*, *TARDBP*, *FUS*, *TBK1*, *C9orf72*) and *APOE* genotyping.

### Neuropathological assessment

For patients with available autopsy data, neuropathological findings were reviewed based on standard neuropathological examination, including immunohistochemical assessment of α-synuclein, TDP-43, amyloid-β, and tau pathology. The distribution and severity of Lewy pathology were classified according to Braak LB stage and the modified McKeith classification,^25–27^ and TDP-43 pathology according to Brettschneider stage.^28^ Amyloid-β deposition was assessed using the Thal phase, neuritic plaque pathology using the CERAD score, and tau pathology using Braak stage with AT8 immunohistochemistry.^29–31^

### Statistical analysis

All the analyses below were based on participants with available follow-up clinical data and were performed in *R* version 4.5.0 using the *mice*, *survival*, *lme4*, and *lmerTest* packages.

Baseline characteristics were compared across αSyn SAA categories using the Kruskal– Wallis rank-sum test for continuous variables and Fisher’s exact test for categorical variables, with pairwise comparisons against the negative group. Kinetic parameters were compared between weak– and strong-positive samples using the Wilcoxon rank-sum test.

We assessed two prognostic outcomes: (i) survival, defined as time to death or tracheostomy, and (ii) longitudinal ALSFRS-R decline. Both were evaluated in unadjusted models and models adjusted for baseline age, ΔFS, %FVC, and NfL, based on previous literature and data availability.^3,32^ In the pooled analyses, survival was assessed using Cox proportional hazards models stratified by cohort, while longitudinal ALSFRS-R scores were assessed using linear mixed-effects (LME) models with participant-specific random intercepts and slopes and cohort as a fixed effect. The LME analyses also evaluated an NfL × αSyn SAA × time interaction to assess whether the association between NfL and ALSFRS-R decline differed by αSyn SAA status. Models for both outcomes were also fitted separately within each cohort to assess the consistency of estimates. Fluid biomarker values were z-scored within each cohort to account for assay differences.

Missing values in variables required for the core adjustment set were handled using multiple imputation with predictive mean matching, generating 30 imputed datasets.^33^ Baseline ALSFRS-R, required to calculate ΔFS, was missing in 2.5% of the pooled cohort, and %FVC in 5.0%; age and CSF NfL had no missing values. ΔFS was calculated after imputation of baseline ALSFRS-R. Imputation was performed in the full analytic sample before subgroup restrictions, and the same imputed datasets were used for the primary and sensitivity analyses; estimates were pooled using Rubin’s rules.^34^

The primary Cox and LME analyses were restricted to participants without evidence of familial or genetically defined ALS, based on available genetic and family history data, because of reported prognostic differences in these forms of ALS.^35^ Sensitivity analyses included all participants with ALS, irrespective of familial or genetic status, or were restricted to Aβ-negative participants. We repeated both analyses using multiple imputation and complete-case approaches.

### Ethics

This study followed the 2024 Declaration of Helsinki. The Nagoya cohort was approved by the Ethics Review Committee of Nagoya University Graduate School of Medicine (2013-0035, 2015-0041, 2022-0155) and complied with Japanese governmental ethical guidelines. All prospectively enrolled participants provided written informed consent; genetic testing was performed only in those who provided separate consent for it. Retrospective use of existing data and specimens followed an approved opt-out procedure publicized on the institutional website. BioFINDER (NCT03174938) and Bologna studies were approved by the Regional Ethical Committee in Lund (2016/1053) and the Ethics Committee of Area Vasta Emilia Centro (CE-AVEC 17151-17152), respectively, and all participants in both studies provided written informed consent.

## Results

### αSyn SAA positivity in the ALS cohorts

The pooled analytic cohort comprised 443 patients with ALS (**Table 1**; Nagoya, *n* = 248, Bologna, *n* = 195). Baseline characteristics were comparable between cohorts, including age (median: 67 in both cohorts), sex (females, 41% vs 38%), %FVC (median: 93% vs 90%), ΔFS (median: 0.46 vs 0.44), and CSF Aβ status (positive, 15% vs 13%; **Supplementary Table S1**). Across both cohorts, 24/443 patients (5.4%) were classified as strong-positive for αSyn SAA. A further 13 patients were classified as weak-positive, yielding an overall αSyn SAA positivity rate of 8.4% (37/443). αSyn SAA classifications were comparable between the Nagoya and Bologna cohorts (strong-positive, 6.5% vs 4.1%; weak-positive, 2.8% vs 3.1%; *P* = 0.57).

**Table 1.** Baseline characteristics by αSyn SAA category.

| Variable | Overall<br>*n* = 443 | Negative<br>*n* = 406 | Weak Positive<br>*n* = 13 | Strong Positive<br>*n* = 24 | *P*-value,<br>weak positive vs negative<sup>a</sup> | *P*-value,<br>strong positive vs negative<sup>a</sup> |
| --- | --- | --- | --- | --- | --- | --- |
| Age at baseline (years) | 67 (58, 73) | 67 (57, 73) | 74 (66, 76) | 68 (65, 74) | 0.041 | 0.077 |
| Sex (Female) | 176 (40%) | 163 (40%) | 5 (38%) | 8 (33%) | >0.9 | 0.67 |
| Onset site |  |  |  |  | 0.74 | 0.63 |
| Bulbar/Respiratory | 108 (24%) | 99 (24%) | 2 (15%) | 7 (29%) |  |  |
| Spinal | 335 (76%) | 307 (76%) | 11 (85%) | 17 (71%) |  |  |
| Pathogenic variant in ALS-associated genes<sup>b</sup> | 35 (9.4%) | 33 (9.7%) | 1 (8.3%) | 1 (5.0%) | >0.9 | 0.71 |
| Disease duration (months) | 13 (8, 24) | 13 (8, 24) | 22 (14, 27) | 12 (7, 16) | 0.037 | 0.37 |
| ΔFS<sup>b</sup> | 0.45 (0.22, 0.89) | 0.44 (0.21, 0.90) | 0.47 (0.29, 0.69) | 0.56 (0.35, 0.89) | 0.92 | 0.19 |
| ALSFRS-R<sup>b</sup> | 42 (39, 44) | 42 (39, 44) | 39 (36, 42) | 41 (40, 44) | 0.050 | 0.55 |
| FVC % predicted<sup>b</sup> | 92 (76, 108) | 92 (77, 108) | 94 (53, 115) | 83 (71, 102) | 0.84 | 0.091 |
| CSF NfL (z-score)<sup>c</sup> | 0.06 (-0.64, 0.70) | 0.06 (-0.66, 0.69) | 0.11 (-0.09, 0.84) | 0.18 (-0.22, 0.87) | 0.44 | 0.46 |
| CSF Aβ status (positive) | 62 (14%) | 54 (13%) | 5 (38%) | 3 (13%) | 0.025 | >0.9 |
| *APOE* ε4 carrier<sup>b</sup> | 61 (18%) | 56 (18%) | 2 (18%) | 3 (16%) | >0.9 | >0.9 |
| MMSE<sup>b</sup> | 29 (27, 30) | 29 (27, 30) | 28 (27, 30) | 29 (26, 29) | 0.79 | 0.65 |
| Cohort (Bologna/Nagoya) | 195/248 (44%/56%) | 181/225 (45%/55%) | 6/7 (46%/54%) | 8/16 (33%/67%) | >0.9 | 0.30 |

*n* (%); Median (Q1, Q3)
<sup>a</sup>Continuous variables: Kruskal-Wallis test. Categorical variables: Fisher's exact test. Weak vs negative *P*-values are exploratory given small sample size (Bologna *n* = 6, Nagoya *n* = 7).
<sup>b</sup>Missing data: genetic status, *n*=66 (negative), *n*=1 (weak positive), *n*=4 (strong positive); ΔFS, *n*=10 (negative), *n*=1 (weak positive); ALSFRS-R, *n*=10 (negative), *n*=1 (weak positive); %FVC, *n*=19 (negative), *n*=2 (weak positive), *n*=1 (strong positive); *APOE* $\epsilon$ 4 carrier status, *n*=91 (negative), *n*=2 (weak positive), *n*=5 (strong positive); MMSE, *n*=57 (negative), *n*=1 (strong positive).
<sup>c</sup>CSF NfL expressed as within-cohort z-score (log-transformed) to harmonize assay differences between cohorts.
**Abbreviations:** Aβ , amyloid-β ; ALS, amyotrophic lateral sclerosis; ALSFRS-R, ALS Functional Rating Scale-Revised; *APOE*, apolipoprotein E; αSyn SAA, α-synuclein seed amplification assay; CSF, cerebrospinal fluid; FVC, forced vital capacity; MMSE, Mini-Mental State Examination; NfL, neurofilament light chain; ΔFS, disease progression rate from symptom onset to baseline.

To contextualize this prevalence, we compared ALS with diagnostic groups across the AD continuum in the BioFINDER cohort (**Fig. 1A**). αSyn SAA positivity increased from 5.5% in Aβ-negative CU participants and 10.8% in Aβ-positive CU participants to 16.5% in Aβ-positive MCI and 21.6% in AD dementia. In age– and sex-adjusted comparisons, ALS did not show significantly higher odds of αSyn SAA positivity than Aβ-negative CU (OR 1.33, 95% CI 0.82–2.14) or Aβ-positive CU (OR 0.91, 0.54–1.56) and showed significantly lower odds than AD dementia (OR 0.44, 0.27–0.70). Aβ status was not significantly associated with αSyn SAA positivity within ALS (OR 1.22, 0.49–2.80; **Supplementary Fig. S1**).

**Fig. 1.**
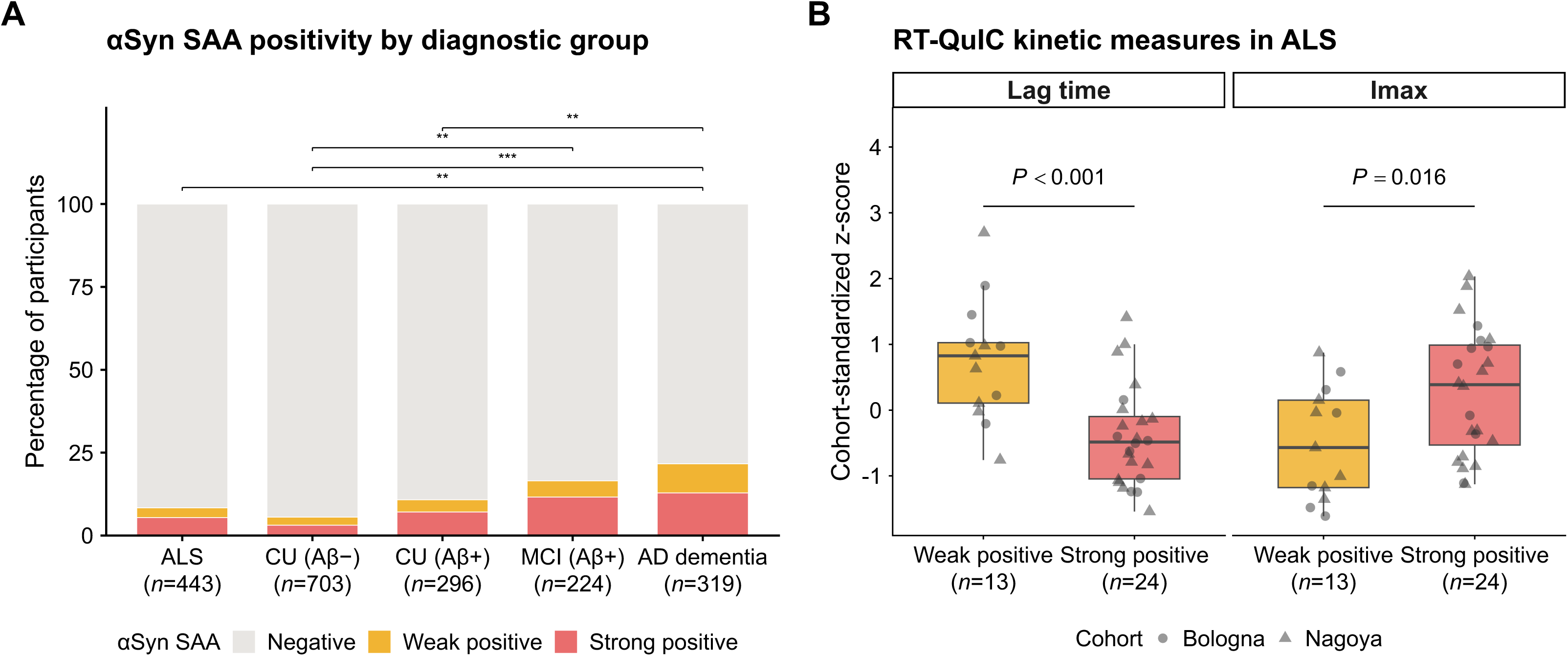
αSyn SAA positivity across diagnostic groups and association with RT-QuIC parameters. (**A**) αSyn SAA negative, weak-positive, and strong-positive samples across ALS, CU (Aβ−), CU (Aβ+), MCI (Aβ+), and AD dementia groups. Asterisks above brackets indicate comparisons of αSyn SAA positivity odds using age– and sex-adjusted logistic regression (*P* < 0.05 *, *P* < 0.01 **, *P* < 0.001 ***, BH-FDR-corrected). (**B**) Cohort-standardized z-scores of RT-QuIC lag time and Imax in weak-positive (*n*=13) versus strong-positive (*n*=24) αSyn SAA samples from patients with ALS (Bologna and Nagoya cohorts). Boxes show median and IQR; *P* values from the Wilcoxon rank-sum test. **Abbreviations:** AD, Alzheimer’s disease; Aβ, amyloid-beta; αSyn SAA, alpha-synuclein seed amplification assay; BH-FDR, Benjamini-Hochberg false discovery rate; CU, cognitively unimpaired; Imax, maximum fluorescence intensity; IQR, interquartile range; MCI, mild cognitive impairment; RT-QuIC, real-time quaking-induced conversion.

### αSyn SAA categories differed in seeding activity and baseline characteristics

Across both ALS cohorts, strong-positive samples showed higher Imax values and shorter lag times, consistent with greater αSyn seeding activity (**Fig. 1B**). Further stratification by the number of positive wells showed a similar pattern, with progressively shorter lag times and higher Imax values as the number of positive wells increased (**Supplementary Fig. S2**).

The strong-positive αSyn SAA subgroup showed no clear baseline demographic or clinical differences from the negative group (**Table 1**). In contrast, the weak-positive αSyn SAA subgroup exhibited a distinct profile compared to the negative subgroup, characterized by higher age (median 74 years; weak vs. negative *P* = 0.041), longer disease duration (median 22 months; weak vs. negative *P* = 0.037), and an elevated prevalence of CSF Aβ positivity (38% vs. 13%; weak vs. negative *P* = 0.025).

### αSyn SAA strong positivity was associated with shorter survival in ALS

In the pooled cohort, the αSyn SAA strong-positive subgroup had significantly shorter tracheostomy-free survival than the negative subgroup (median 14 vs. 31 months; log-rank *P* = 0.019; **Fig. 2A**). This association remained significant after adjustment for age, ΔFS, %FVC, and NfL, with directionally consistent estimates in each cohort (**Fig. 2B**). Sensitivity analyses that included patients with a family history of ALS or known ALS-associated mutations, or were restricted to Aβ-negative patients, also yielded consistent prognostic associations (**Fig. 2B**; **Supplementary Fig. S3**). In contrast, no significant survival differences were observed between the αSyn SAA weak-positive and negative subgroups or between the Aβ-positive and Aβ-negative subgroups.

**Fig. 2.**
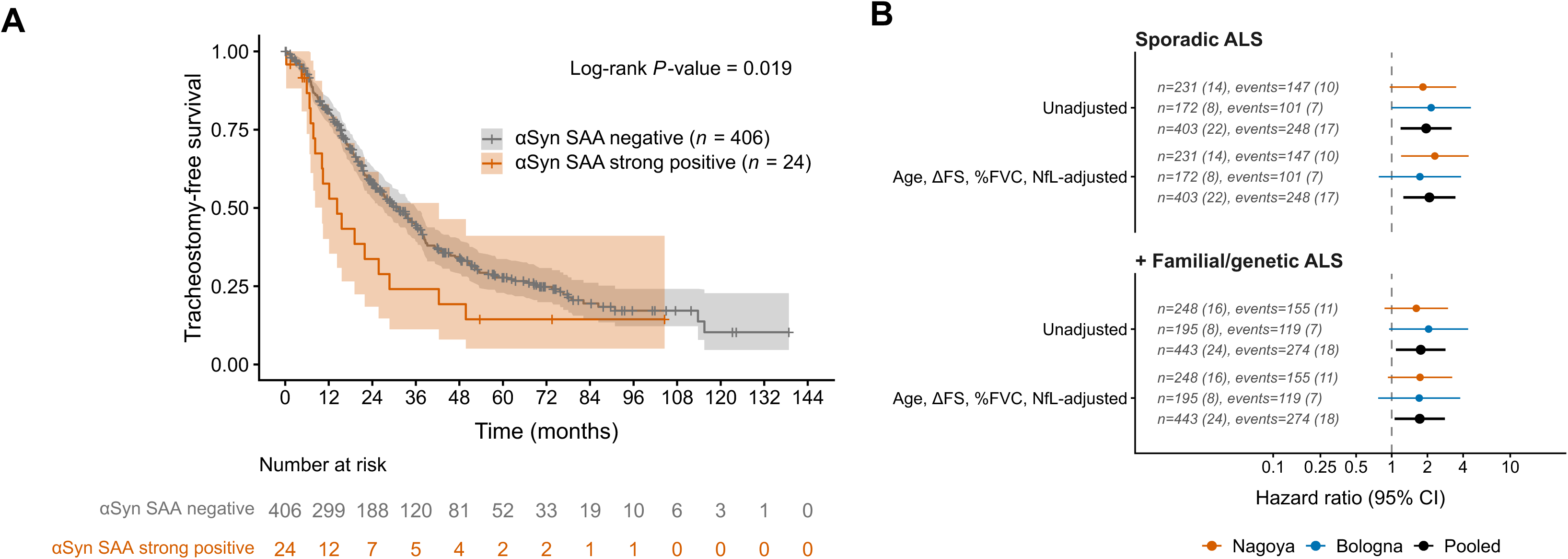
Survival analyses for αSyn SAA strong positivity. **(A)** Kaplan-Meier curves for time to death or tracheostomy in the pooled cohort, comparing αSyn SAA strong-positive (*n* = 24) and negative (*n* = 406) subgroups. **(B)** Cox hazard ratios for αSyn SAA strong positivity (unadjusted; age, ΔFS, %FVC, NfL-adjusted). Primary analysis (sporadic ALS) and sensitivity analysis adding familial and genetic ALS. Estimates are shown for each cohort and for the pooled cohort. Horizontal lines indicate 95% confidence intervals; numbers of patients and events (strong-positive in parentheses) are shown for each estimate. **Abbreviations:** ΔFS, disease progression rate from symptom onset to baseline; αSyn SAA, alpha-synuclein seed amplification assay; NfL, neurofilament light; %FVC, percent predicted forced vital capacity.

### αSyn SAA modified the relationship between NfL and ALSFRS-R decline

Although strong αSyn SAA positivity was associated with shorter survival, its association with faster ALSFRS-R decline did not reach statistical significance in the LME models (αSyn SAA × time: β = −0.24, *P* = 0.16). Estimates were nevertheless generally in the direction of faster decline in strong-positive patients across all sensitivity analyses (**Supplementary Fig. S4A**). Note that patients with aggressive disease courses who reached the survival endpoint early contributed fewer ALSFRS-R observations to the LME models.

We then examined whether the association between NfL and ALSFRS-R decline differed by αSyn SAA status. The NfL × strong αSyn SAA positivity × time interaction was significant in both unadjusted models and models adjusted for age, ΔFS, and %FVC (**Fig. 3**). In cohort-specific analyses using the adjusted model, estimates were directionally consistent (Nagoya, β = −0.71, *P* = 0.034; Bologna, β = −0.95, *P* = 0.27), although the smaller number of strong-positive cases in Bologna yielded a less precise estimate (**Supplementary Fig. S4B**).

**Fig. 3.**
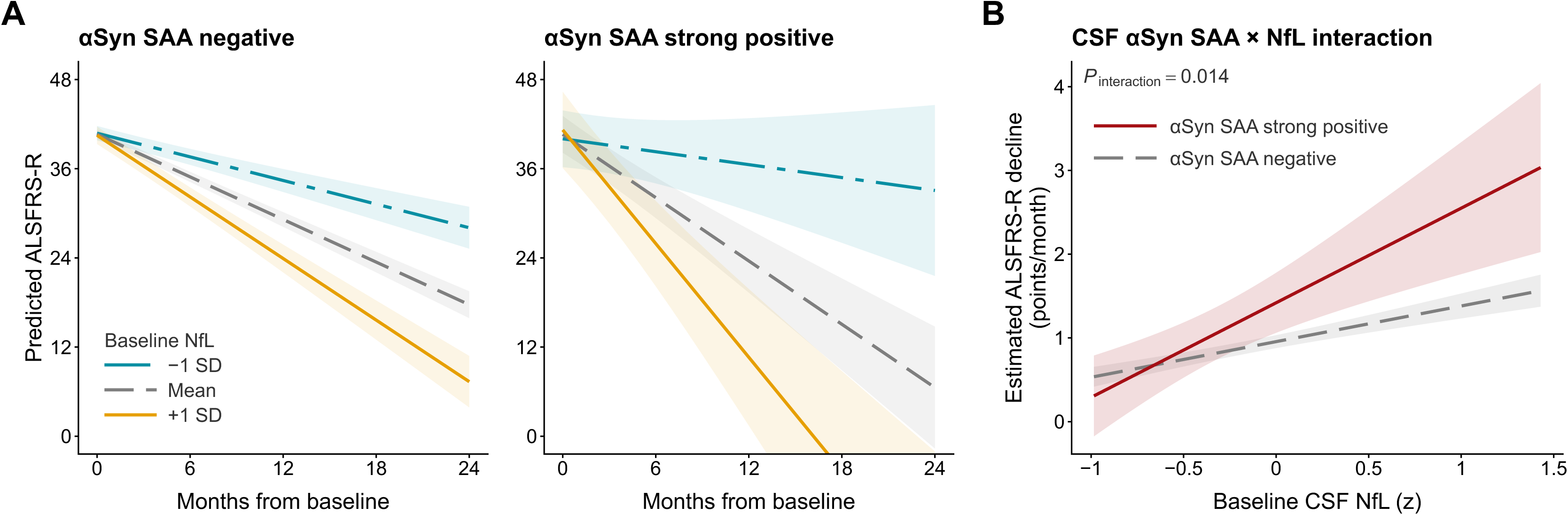
Interaction between baseline CSF NfL and αSyn SAA status on longitudinal ALSFRS-R decline. **(A)** Predicted ALSFRS-R trajectories over 24 months from pooled core covariates-adjusted linear mixed-effects models, stratified by CSF αSyn SAA status, shown at baseline CSF NfL levels of −1 SD, mean, and +1 SD. **(B)** Estimated monthly ALSFRS-R decline rate (points/month) as a continuous function of baseline CSF NfL z-score, from the same models, stratified by αSyn SAA strong positivity status. Shaded bands indicate 95% confidence intervals. Interaction P-value corresponds to the αSyn SAA strong positivity × NfL × time term. **Abbreviations:** ALSFRS-R, ALS Functional Rating Scale-Revised; αSyn SAA, alpha-synuclein seed amplification assay; CSF, cerebrospinal fluid; NfL, neurofilament light; SD, standard deviation.

The NfL × Aβ positivity × time interaction was also significant (**Supplementary Fig. S5**). Notably, both interactions remained significant in adjusted models after excluding patients positive for the other biomarker: the αSyn SAA interaction among Aβ-negative patients (**Supplementary Fig. S4B**) and the Aβ interaction among αSyn SAA-negative patients (**Supplementary Fig. S5**). These findings were preserved when familial and genetic ALS cases were included (**Supplementary Figs. S4B and S5**).

### Neuropathological findings supported the CSF αSyn SAA results

Neuropathological data were available for five patients, including one αSyn SAA-strong positive and four αSyn SAA-negative cases, all from the Nagoya cohort. All five had neuropathologically confirmed ALS; one patient had *FUS*-associated ALS, while the remaining four had sporadic ALS with TDP-43 pathology (**Supplementary Table S3**).

The case with strong αSyn SAA positivity had widespread TDP-43 pathology (Brettschneider stage 4) and Bunina bodies, together with extensive Lewy pathology (Braak LB stage 6; diffuse neocortical pattern; **Fig. 4**). No parkinsonism or autonomic dysfunction had been documented in the medical records during life. Among the four αSyn SAA-negative cases, no Lewy pathology was detected in three, whereas one showed only sparse Lewy pathology restricted to the amygdala, classified as an amygdala-predominant pattern (**Supplementary Table S3**).

**Fig. 4.**
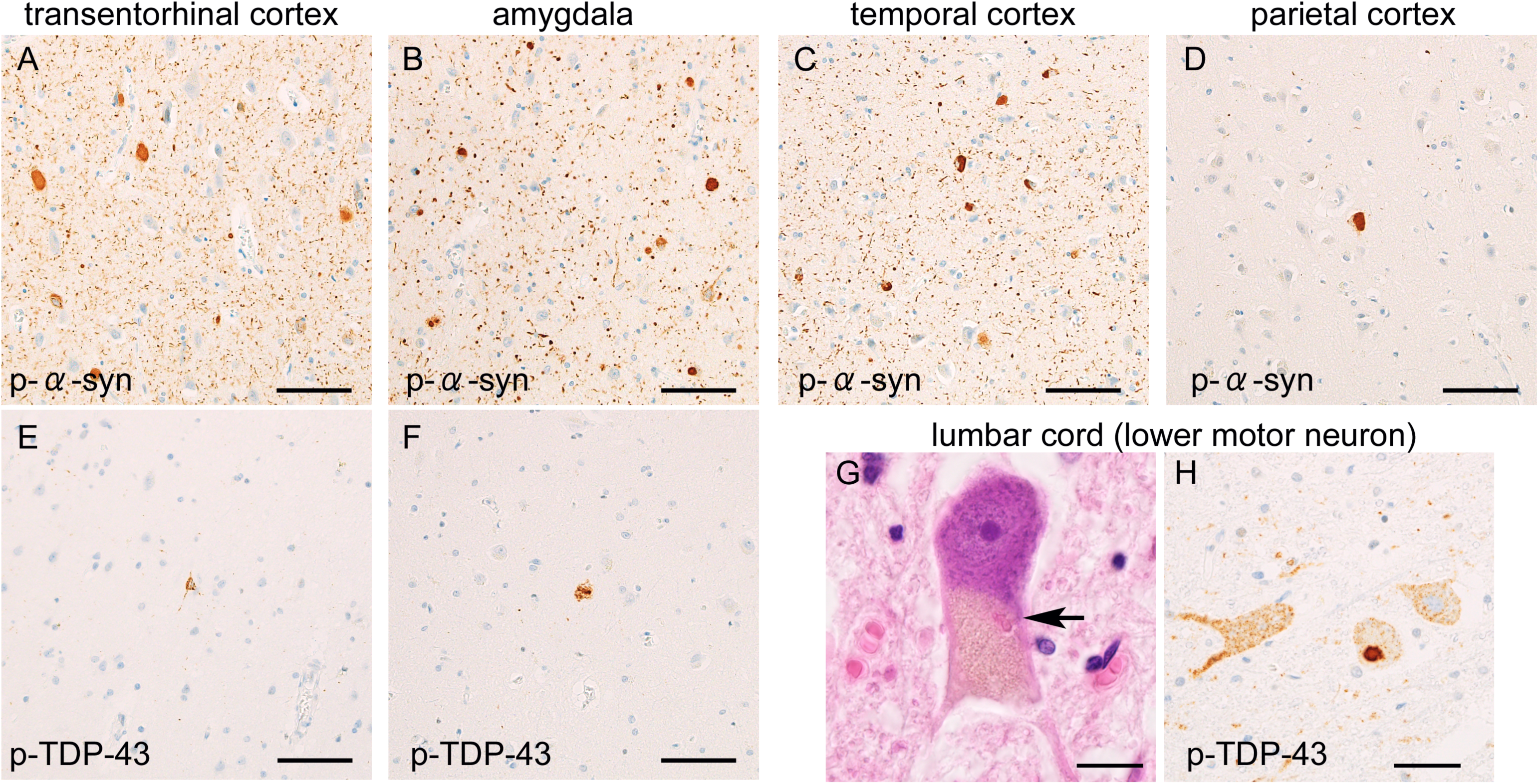
Combined α-synuclein and TDP-43 pathology in an autopsied case. A postmortem assessment revealed marked α-synuclein aggregation involving the transentorhinal cortex **(A)**, amygdala **(B)**, and neocortices (**C and D**: temporal and parietal cortices are shown). The case also showed mild TDP-43 aggregates in neurons of the transentorhinal cortex (**E**) and amygdala (**F**), along with Bunina bodies (**G**, arrow) and TDP-43 aggregates (**H**) within the lower motor neurons, diagnostic of ALS-TDP. **(A–D)** Anti-phosphorylated α-synuclein immunohistochemistry **(E, F, and H),** anti-phosphorylated TDP-43 immunohistochemistry (G), and hematoxylin-eosin staining (A-D). Scale bars = (A–F) 50 μm and (G–H) 10 μm. **Abbreviations:** ALS, amyotrophic lateral sclerosis; ALS-TDP, ALS with TDP-43 pathology; TDP-43, TAR DNA-binding protein 43.

## Discussion

Although infrequent, strong αSyn SAA positivity delineated a subgroup with a distinct prognostic profile across two independent ALS cohorts. Strong positivity on this assay was associated with significantly shorter survival and appeared to modify the relationship between CSF NfL and functional deterioration.

Previous neuropathological investigations have reported α-synuclein pathology in approximately 11% of ALS cases,^5^ which aligns with the overall positivity rate (including weak-positives, ∼8.4%) observed in the current study. While a prior study using post-mortem CSF αSyn SAA demonstrated a higher prevalence of 14%,^9^ direct comparisons are precluded by variations in biospecimen characteristics and assay protocols. The higher proportion in post-mortem samples may be partially attributable to the advanced age and neurodegenerative stage relative to the time of baseline CSF collection in our cohorts. In our analyses, the odds of αSyn SAA positivity in ALS were not significantly higher than in Aβ-negative CU individuals and were lower than in AD dementia, after adjustment for age and sex. Although genetic and transcriptomic studies have suggested modest overlap between ALS and Parkinson’s disease,^36,37^ and the soluble prion-like domain of TDP-43 and α-synuclein has been shown to interact and form hybrid fibrils *in vitro*,^38^ our findings did not support an ALS-specific predisposition to αSyn SAA positivity. The observed positivity rate was consistent with the reported prevalence of incidental Lewy body pathology in older individuals (approximately 8–15%),^13,39,40^ suggesting largely age-related co-pathology; this warrants neuropathological validation in larger ALS cohorts with age-matched controls.

Based on the established analytical protocols,^21^ we stratified participants into strong-versus weak-positivity. Reactivity across three or four wells on initial testing indicates robust α-synuclein seeding activity, consistent with a higher seed burden.^14^ Weak positivity, established only after repeat testing, reflects lower seed concentrations, presumably associated with a lower burden or more restricted distribution of Lewy body co-pathology.^13,14^ In our ALS cohort, lag time and maximum fluorescence aligned with replicate reactivity, supporting this distinction. Neuropathological findings provided additional, albeit exploratory, support for the interpretation of these results. While autopsy data were available for only one case with strong αSyn SAA positivity and four with negative αSyn SAA results, the observed pattern was consistent with previous evidence that CSF αSyn SAA more reliably detects widespread than focal, low-burden Lewy pathology,^13,41^ a correspondence documented so far mainly in cohorts other than ALS.

Notably, the weak-positive subgroup showed a distinct baseline profile, characterized by older age, longer disease duration at CSF sampling, and a higher prevalence of Aβ positivity, despite a ΔFS comparable to that of SAA-negative patients. This pattern raises the possibility that the weak-positive subgroup comprises patients who had already survived longer before sampling and accumulated age-related or incidental co-pathology,^7^ potentially reflecting survivorship-related enrichment. In contrast, this pattern was not observed in the strong-positive subgroup, which had a disease duration comparable to that of SAA-negative patients but substantially shorter subsequent survival. Although the small number of weak-positive cases precludes firm conclusions, these findings support considering weak and strong αSyn SAA positivity separately rather than combining them into a single positive category.

For the prognostication of ALS, the observed interaction between CSF NfL and co-pathologies, including both Aβ and αSyn SAA strong positivity, suggests that these comorbid proteinopathies do not exert a uniform, independent effect on functional progression. Instead, they appear to specifically amplify functional loss when baseline neurodegenerative activity is high, as indicated by elevated NfL. This is plausible if low NfL is taken to reflect a correspondingly low level of ALS-related neuroaxonal damage: under such conditions, co-existing Aβ or α-synuclein pathology alone may not produce a detectable effect on ALSFRS-R decline, whereas their modifying effect may become more apparent in the presence of greater ongoing neurodegenerative activity.

Interestingly, αSyn SAA strong positivity was additionally associated with shorter survival after adjustment for NfL and core clinical predictors, whereas Aβ positivity showed no comparable association. Aβ may primarily modify the prognostic effect of NfL, whereas α-synuclein pathology may also have an independent association with survival. Hypothetically, α-synuclein-related pathology may affect survival through complications not fully captured by the ALSFRS-R trajectory, such as non-motor symptoms. Another possibility, albeit unlikely, is that a few individuals in the αSyn SAA strong-positive group had a primary synucleinopathy that mimicked ALS. Although such misclassification cannot be entirely excluded, primary synucleinopathies generally have longer survival than typical ALS. Their inclusion would therefore be expected to attenuate, rather than generate, the association between strong positivity and poorer survival. The observed adverse prognostic association may thus be conservative.

Identifying these rare but prognostically distinct patient subgroups has implications for both patient communication and clinical trial design in ALS. Incorporating CSF Aβ42/40 and αSyn SAA status alongside NfL may permit more precise stratification of disease progression trajectories. However, clinical features alone may not provide sufficient guidance on which patients should undergo CSF testing for these co-pathology assessments. Prior neuropathological studies of ALS with concomitant Lewy body pathology have demonstrated heterogeneous clinical manifestations, with parkinsonism and associated non-motor symptoms frequently remaining clinically occult.^6,42^ Consistent with this, the strong-positive αSyn SAA subgroup in our cohorts did not exhibit a distinct pattern of baseline clinical characteristics compared with the negative subgroup. Although αSyn SAA positivity may underlie atypical ALS presentations that cannot be fully explained by upper or lower motor neuron involvement, parkinsonism may be particularly difficult to assess in ALS, especially at later stages, when muscle atrophy and profound weakness may mask it.^42^

In clinical trials, adjusting for established prognostic baseline factors is recommended to improve statistical efficiency and treatment-effect estimation.^43^ NfL is particularly attractive for this purpose in ALS because of its strong and reproducible prognostic value.^3,10,44^ However, adjusting for NfL alone may not fully capture prognostic heterogeneity if its association with subsequent functional decline varies by underlying co-pathology. Given the low prevalence of these co-pathologies and their potentially complex biological interactions, a pragmatic initial strategy might be to enrich for a more biologically homogeneous ALS population without evidence of AD-or Lewy body co-pathology, particularly in trials where reducing prognostic heterogeneity is a priority. Future studies could further explore whether, rather than excluding these subgroups, co-pathology status and its modifying effects on established prognostic markers may be worth incorporating into trial designs. Ultimately, this may also raise the possibility that therapies targeting AD-or Lewy body-pathology could modify disease trajectories in co-pathology-positive subsets of ALS.

Several limitations merit consideration. First, the low prevalence of αSyn SAA positivity limited statistical power, and our stratified prognostic analyses in ALS require independent validation despite consistent sensitivity analyses. Although strong/weak-positive classification was derived from the assay’s established *a priori* positivity criteria,^21^ its application to prognostic stratification remains to be validated. The small number of patients positive for both Aβ and αSyn SAA also precluded assessment of their interactive effects on prognosis. Second, the study lacked systematic clinical assessment of Lewy body disease-related features, dopamine transporter imaging, and neuropathological confirmation in most participants, limiting our ability to determine whether αSyn SAA positivity corresponded to clinically manifest or subclinical co-pathology.

## Conclusion

A high α-synuclein seeding profile was uncommon in ALS but identified a prognostically distinct subgroup characterized by shorter survival, with a stronger association between NfL and functional decline. Co-pathology assessment *in vivo* may help elucidate prognostic heterogeneity in ALS and warrants validation for biomarker-based stratification in larger cohorts.

## Data availability

De-identified participant-level clinical data may be shared upon reasonable request to the corresponding authors, in accordance with institutional and legal regulations on patient data confidentiality and applicable data transfer agreements.

## Acknowledgements

We gratefully acknowledge all study participants and their families for their invaluable contributions. We thank all the members of the Department of Neurology, Nagoya University Graduate School of Medicine, for their support on the project. We are grateful to Pilar Lorenzo at Lund University for technical support. We also thank all staff at Minimal Co., Ltd. for the technical support at Nagoya University.

Genetic testing for the Nagoya ALS cohort was performed as part of the Japanese Consortium for Amyotrophic Lateral Sclerosis research (JaCALS) project and the Japan Leading Project for Rare Disease Whole-Genome Sequencing (RDWGS). JaCALS was supported by the Japan Agency for Medical Research and Development (AMED; grant numbers 22wm0425009, 22ek0109492, 24ek0109617, 25ak0101216, and 25wm0625519); Grants-in-Aid for Scientific Research (KAKENHI) from the Japan Society for the Promotion of Science (JSPS; grant numbers 21H05278, 22K07359, 22K07509, 22H02988, 22H04923, 22H03350, 23K06835, 23K06975, 23K24249, and 25K10781); Health and Labour Sciences Research Grants from the Ministry of Health, Labour and Welfare of Japan (grant number 23FC0201); the Hori Sciences and Arts Foundation; and the SERIKA FUND. RDWGS was supported by AMED (grant numbers 23ek0109678 and 23ek0109617).

KO was supported by a fellowship of the Nagoya University CIBoG WISE program from MEXT, THERS Make New Standards Program (funded by JST SPRING, Grant Number JPMJSP2125), and the Industry-University Collaborative Project for Human Resource Development to Accelerate AI R&D in the Health and Medical Fields (MEXT).

ChatGPT (OpenAI, GPT-5.6) and Claude (Anthropic, Sonnet 5 and Opus 5) were used to debug R code and proofread language. No AI tools were used to independently perform data analysis, generate original scientific findings, or draw scientific conclusions. The authors reviewed and confirmed the validity of all content and take full responsibility for the publication.

