## Supplementary for "Cerebrospinal fluid α-synuclein seed amplification assay positivity identifies a prognostically distinct subgroup in amyotrophic lateral sclerosis"

1. **Supplementary Tables** ........................................................................ 2–4
2. **Supplementary Figures** ...................................................................... 5–9

### Supplementary Tables

#### Supplementary Table S1. Clinical characteristics of the Nagoya and Bologna ALS cohorts

| Variable | Nagoya<br>$N = 248$ | Bologna<br>$N = 195$ | *P*-value |
|---|---|---|---|
| Age at baseline (years) | 67 (58, 73) | 67 (59, 74) | 0.46 |
| Sex (Female) | 102 (41%) | 74 (38%) | 0.56 |
| Onset site (Bulbar/Respiratory) | 60 (24%) | 48 (25%) | >0.99 |
| Pathogenic variant in ALS-associated genes | 12 (6.5%) | 23 (12%) | 0.075 |
| Disease duration (months) | 12 (8, 21) | 14 (9, 26) | 0.058 |
| ΔFS | 0.46 (0.25, 0.86) | 0.44 (0.20, 0.94) | 0.55 |
| ALSFRS-R total | 42 (39, 44) | 42 (39, 44) | 0.59 |
| FVC % predicted | 93 (77, 109) | 90 (75, 106) | 0.28 |
| CSF NfL (pg/mL)[a] | 4,102 (2,159, 7,188) | 6,093 (3,011, 10,877) | |
| CSF Aβ status (positive) | 36 (15%) | 26 (13%) | 0.78 |
| CSF αSyn SAA category | | | 0.57 |
| negative | 225 (91%) | 181 (93%) | |
| weak positive | 7 (2.8%) | 6 (3.1%) | |
| strong positive | 16 (6.5%) | 8 (4.1%) | |
| *APOE* ε4 carrier | 36 (20%) | 25 (16%) | 0.40 |
| MMSE | 28 (26, 29) | 29 (28, 30) | <0.001 |
| Follow-up duration (months)[b] | 24 (12, 44) | 19 (9, 34) | 0.002 |
| Event (Death or tracheostomy) | 155 (63%) | 119 (61%) | 0.77 |

Median (Q1, Q3); n (%). Kruskal-Wallis rank sum test; Fisher's exact test.
[a] CSF NfL shown as raw values (pg/mL) for descriptive purposes only; Nagoya and Bologna used different assay platforms, so this raw-value comparison should not be interpreted as a direct biological comparison. All formal analyses use within-cohort z-scores to harmonize assay differences.
[b] Follow-up duration is shown alongside the event rate (death or tracheostomy). Note that differences in follow-up length between cohorts (e.g. data cut-off dates) can confound comparisons that do not account for censoring.
Missing values (Nagoya, Bologna): pathogenic variant status (*n*=64, *n*=7); ΔFS (*n*=8, *n*=3); ALSFRS-R total (*n*=8, *n*=3); FVC % predicted (*n*=1, *n*=21); APOE4 carrier status (*n*=64, *n*=34); MMSE (*n*=10, *n*=48).
**Abbreviations:** ALSFRS-R, ALS Functional Rating Scale-Revised; APOE, apolipoprotein E; Aβ, amyloid-beta; αSyn SAA, alpha-synuclein seed amplification assay; CSF, cerebrospinal fluid; ΔFS, disease progression rate from symptom onset to baseline; FVC, forced vital capacity; MMSE, Mini-Mental State Examination; NfL, neurofilament light; Q1, first quartile; Q3, third quartile.

#### Supplementary Table S2. Demographic and clinical characteristics stratified by diagnosis and Aβ status

| Characteristic | ALS (Aβ−) *n* = 381 | ALS (Aβ+) *n* = 62 | CU (Aβ−) *n* = 703 | CU (Aβ+) *n* = 296 | MCI (Aβ+) *n* = 224 | AD dementia *n* = 319 |
|---|---|---|---|---|---|---|
| **Age, years** | 66 (56–72) | 74 (70–79) | 62 (53–75) | 74 (66–79) | 74 (69–77) | 76 (71–79) |
| **Sex** | | | | | | |
| Male | 233 (61%) | 34 (55%) | 305 (43%) | 139 (47%) | 111 (50%) | 136 (43%) |
| Female | 148 (39%) | 28 (45%) | 398 (57%) | 157 (53%) | 113 (50%) | 183 (57%) |
| **MMSE** | 29 (27–30)[a] | 28 (24–29)[b] | 29 (28–30) | 29 (28–30) | 27 (25–28) | 21 (18–24) |
| **αSyn SAA status** | | | | | | |
| negative | 352 (92%) | 54 (87%) | 664 (94%) | 264 (89%) | 187 (83%) | 250 (78%) |
| weak positive | 8 (2.1%) | 5 (8.1%) | 17 (2.4%) | 11 (3.7%) | 11 (4.9%) | 28 (8.8%) |
| strong positive | 21 (5.5%) | 3 (4.8%) | 22 (3.1%) | 21 (7.1%) | 26 (12%) | 41 (13%) |

Median (IQR) or *n* (%).

[a]Missing values: *n*=49.
[b]Missing values: *n*=9.

**Abbreviations:** AD, Alzheimer's disease; Aβ, amyloid-beta; ALS, amyotrophic lateral sclerosis; αSyn SAA, alpha-synuclein seed amplification assay; CU, cognitively unimpaired; IQR, interquartile range; MCI, mild cognitive impairment; MMSE, Mini-Mental State Examination.

#### Supplementary Table S3. Neuropathological assessments

| Case | Age at CSF collection | Duration between CSF collection and death | Sex | CSF αSyn SAA positivity | LB distribution | LB-Braak stage | LB-McKeith | TDP-Brettschneider stage | Aβ Thal phase/CERAD/ Tau-Braak stage (AT8) | Genetic status | CSF NfL (pg/mL) | CSF Aβ status |
|---|---|---|---|---|---|---|---|---|---|---|---|---|
| A | 65–70 | 1–2 years | M | Strong positive | Widespread LB pathology involving brainstem, limbic, and neocortical regions. | 6 | Diffuse neocortical | 4 | 1/0/III | not performed (no consent) | 1,600 | Negative |
| B | 70–75 | < 1 year | F | Negative | Sparse LB pathology, predominantly involving the amygdala. | N/A[a] | Amygdala-predominant | 4 | 1/A/III | not performed (no consent) | 7,300 | Negative |
| C | 70–75 | 1–2 years | M | Negative | None | 0 | 0 | 4 | 0/0/III | no known pathogenic variant | 5,700 | Negative |
| D | 35–40 | < 1 year | M | Negative | None | 0 | 0 | 1 | 0/0/I | no known pathogenic variant | 1,400 | Negative |
| E | 75–80 | 1–2 years | M | Negative | None | 0 | 0 | 0 | 2/A/I | *FUS* | 6,100 | Negative |

[a] LB Braak stage was not assigned because Lewy-related pathology was restricted to the amygdala (amygdala-predominant pattern).
**Abbreviations:** Aβ, amyloid-beta; αSyn SAA, alpha-synuclein seed amplification assay; CERAD, Consortium to Establish a Registry for Alzheimer's Disease; CSF, cerebrospinal fluid; FUS, fused in sarcoma gene; LB, Lewy body; LB-McKeith, McKeith diagnostic criteria for Lewy body disease; NfL, neurofilament light; TDP43, TAR DNA-binding protein 43.

### Supplementary Figures

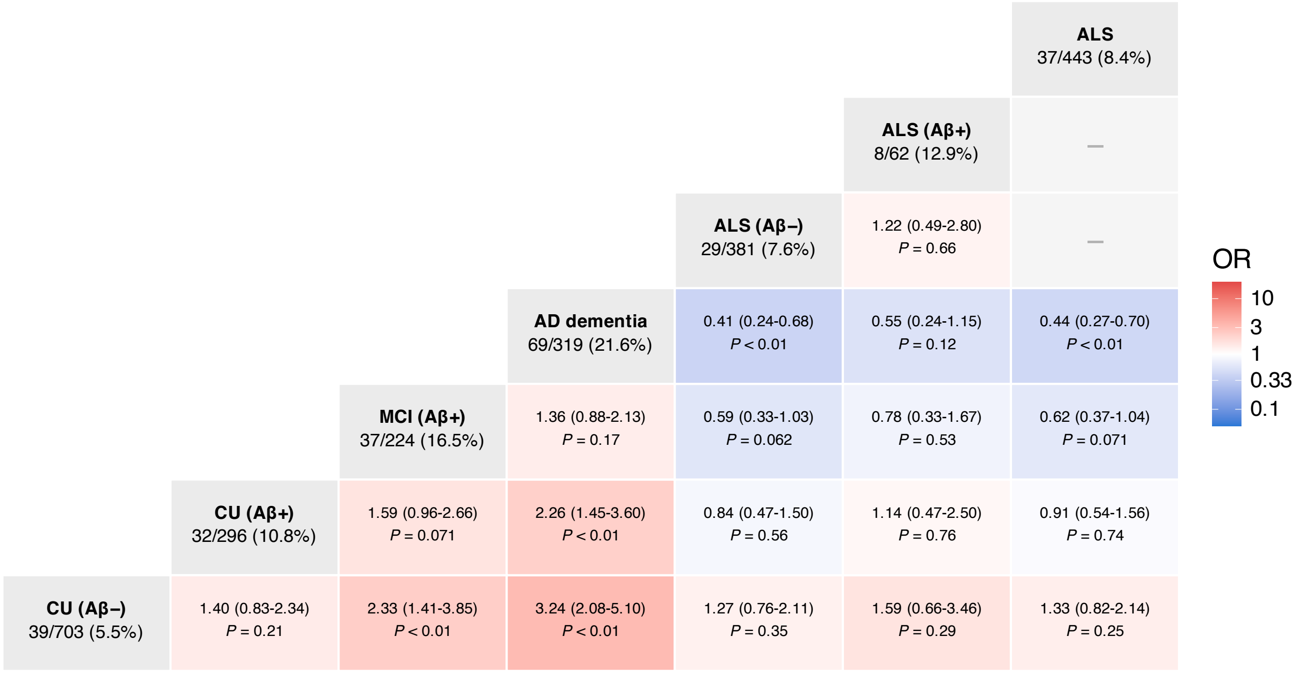


**Supplementary Fig. S1. Age- and sex-adjusted odds of αSyn SAA positivity across diagnostic groups**

Each cell shows the odds ratio (OR; 95% CI, *P*-value) for αSyn SAA positivity in the column group relative to the row group, estimated by logistic regression adjusted for age and sex. Group size and the number of αSyn SAA-positive cases are given in the headers. The rightmost column represents the ALS cohort pooled across Aβ status; dashes denote comparisons not performed because the groups overlap. Cell colour reflects the OR on a logarithmic scale (red, higher odds in the column group; blue, higher odds in the row group). **Abbreviations:** Aβ, amyloid-β; AD, Alzheimer's disease; ALS, amyotrophic lateral sclerosis; CI, confidence interval; CU, cognitively unimpaired; MCI, mild cognitive impairment; OR, odds ratio; αSyn SAA, α-synuclein seed amplification assay.

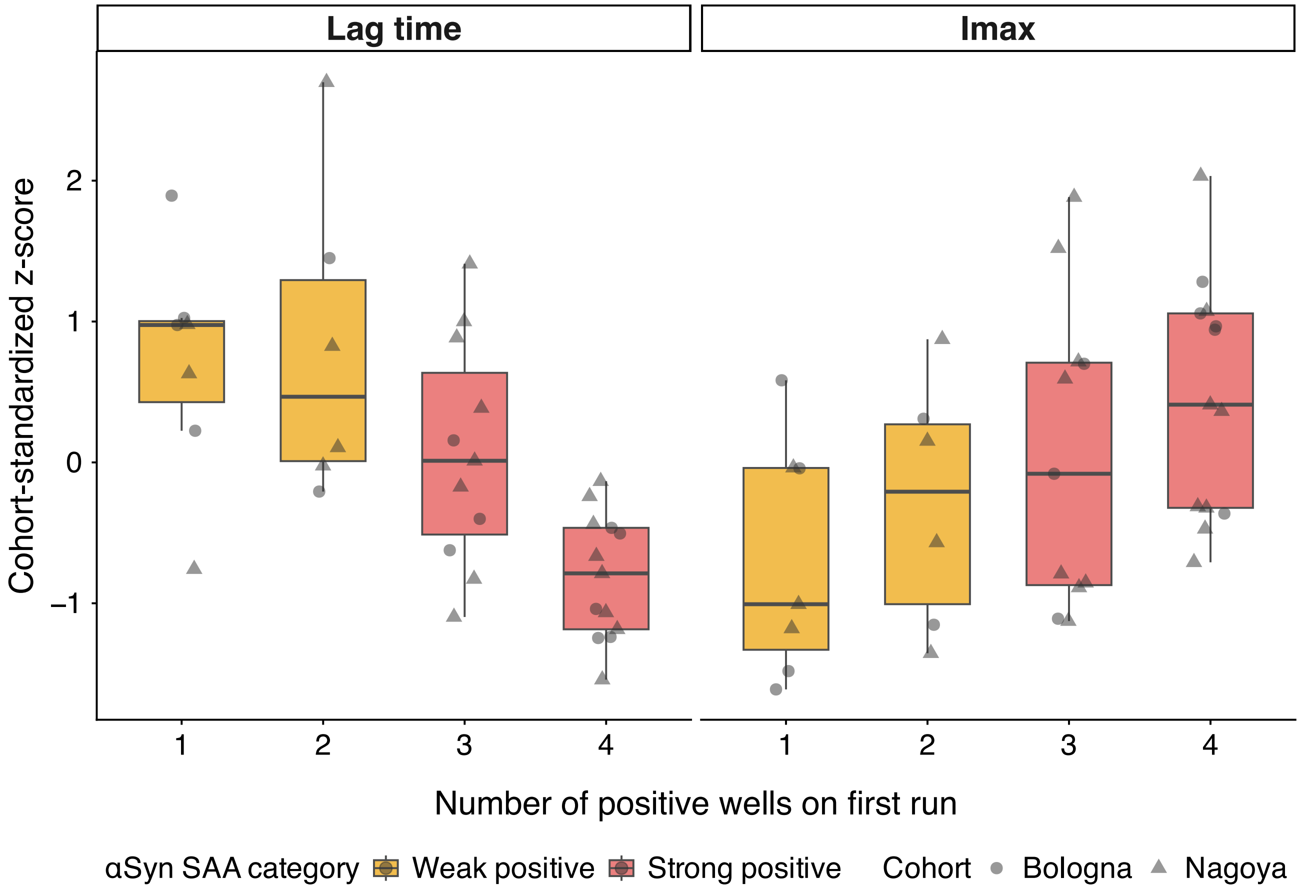


**Supplementary Fig. S2. RT-QuIC kinetics by number of positive wells**

Lag time and maximum fluorescence intensity (Imax) are shown as cohort-standardized z-scores for αSyn SAA-positive ALS patients, stratified by the number of reactive wells in the initial run. Individual patients are overlaid as points. Box colour denotes the αSyn SAA category (orange, weak positive; red, strong positive), and point shape denotes the cohort (circles, Bologna; triangles, Nagoya). **Abbreviations:** ALS, amyotrophic lateral sclerosis; Imax, maximum fluorescence intensity; RT-QuIC, real-time quaking-induced conversion; αSyn SAA, α-synuclein seed amplification assay.

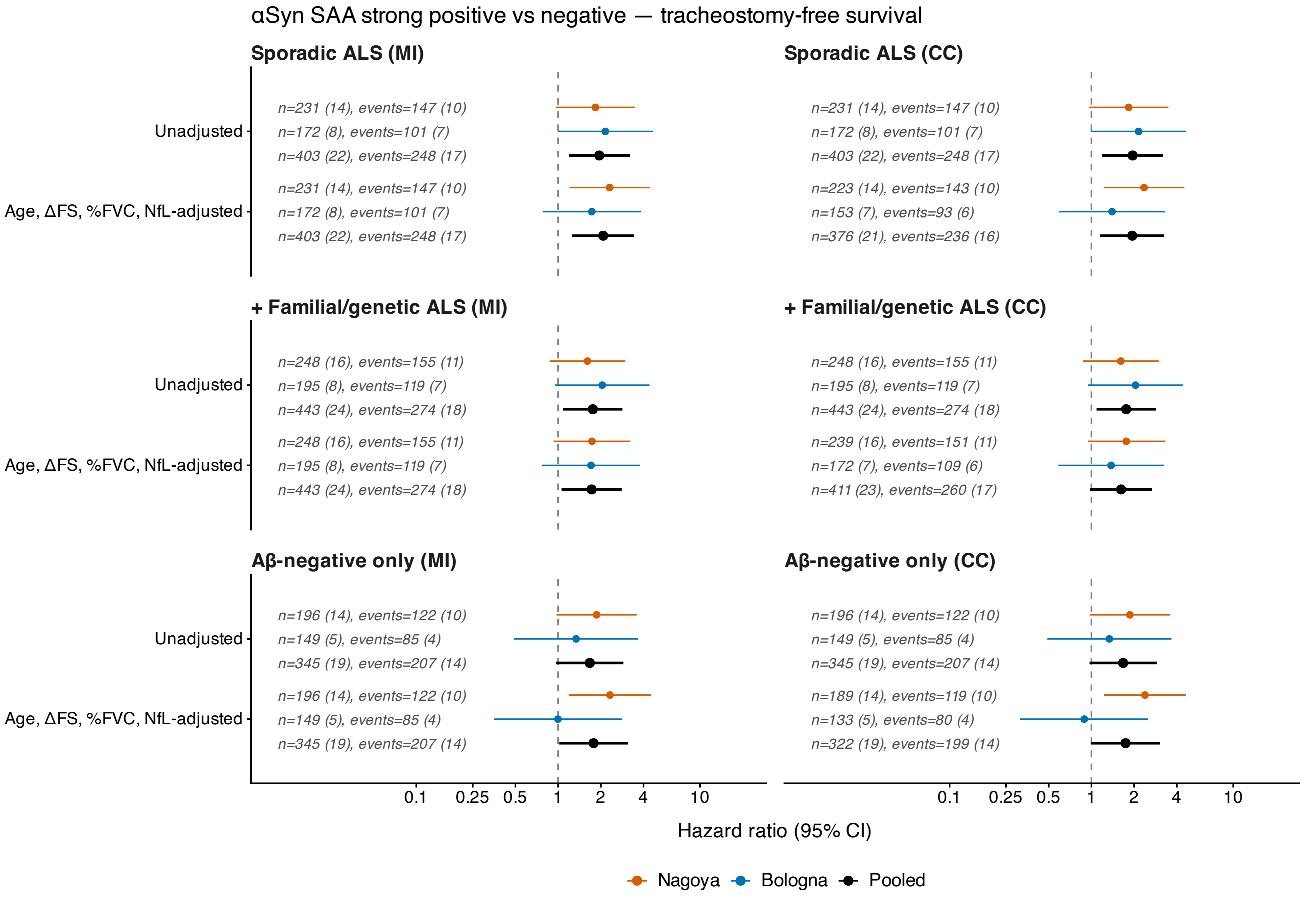


**Supplementary Fig. S3. Sensitivity analyses of Cox models for αSyn SAA status and tracheostomy-free survival**

Hazard ratios (95% confidence intervals) for αSyn SAA strong positive versus negative status on tracheostomy-free survival. Results are presented in the following order: Sporadic ALS (primary analysis), adding familial and genetic ALS, and Aβ-negative only restriction on primary analysis. Each is shown with multiple imputation (MI, left) and complete-case analysis (CC, right), unadjusted and adjusted for age, ΔFS, %FVC, and NfL. Horizontal lines indicate 95% confidence intervals; numbers of patients and events (strong-positive in parentheses) are shown for each estimate. **Abbreviations:** ALSFRS-R, ALS Functional Rating Scale-Revised; Aβ, amyloid-beta; αSyn SAA, alpha-synuclein seed amplification assay; CC, complete case; ΔFS, disease progression rate from symptom onset to baseline; MI, multiple imputation; NfL, neurofilament light; %FVC, percent predicted forced vital capacity.

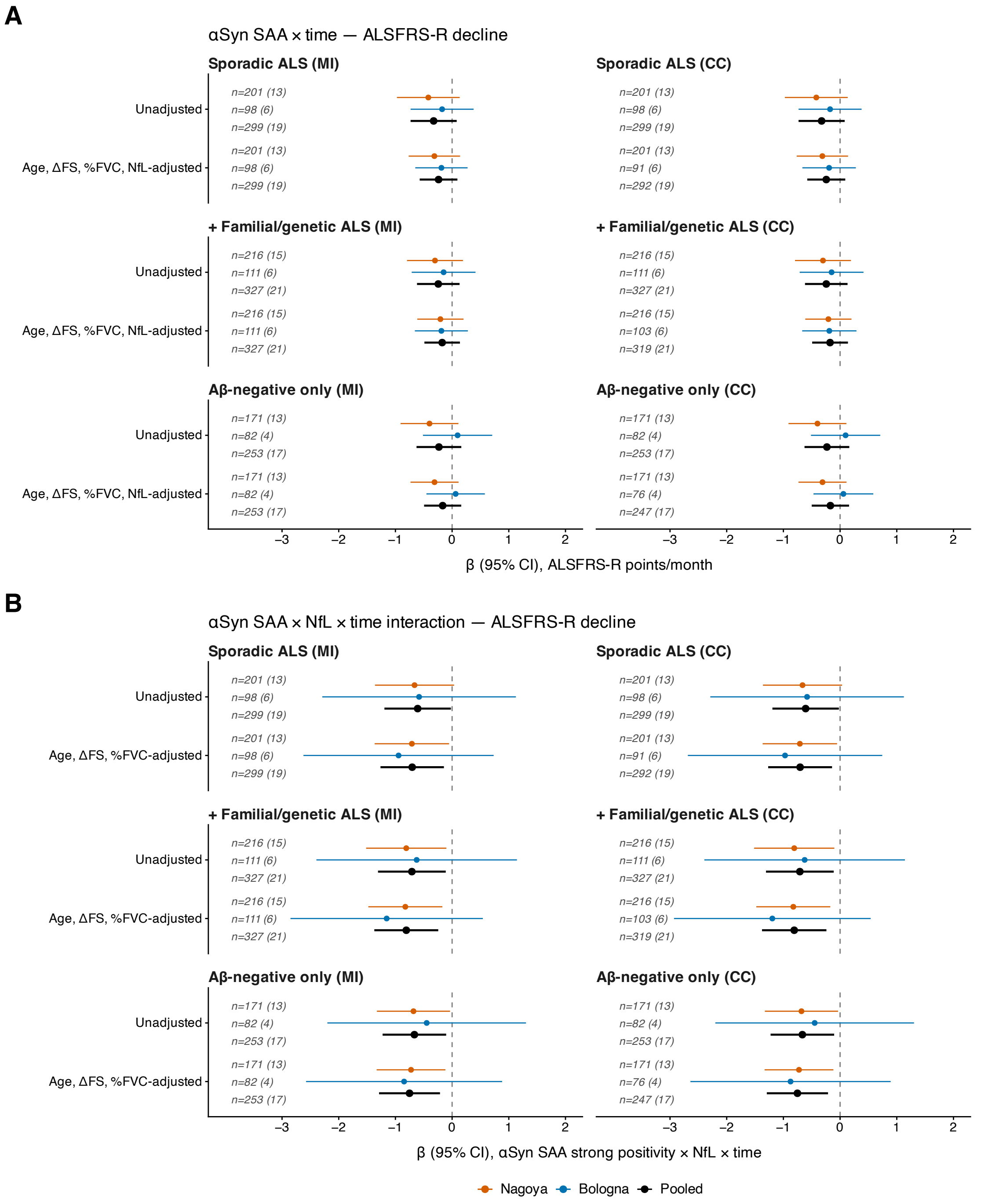


#### Supplementary Fig. S4. Sensitivity analyses of LME models for αSyn SAA status and ALSFRS-R decline

Regression coefficients for αSyn SAA strong-positive versus negative status from linear mixed-effects models, shown for each cohort and for the pooled cohort. **(A)** αSyn SAA × time (difference in ALSFRS-R slope, points/month), unadjusted and adjusted for age, ΔFS, %FVC, and NfL. **(B)** αSyn SAA × NfL × time interaction (points/month per SD of NfL), unadjusted and adjusted for age, ΔFS, and %FVC. Results are presented in the following order: Sporadic ALS (primary analysis), adding familial and genetic ALS, and Aβ-negative only restriction on primary analysis, each with MI (left) and CC (right). Horizontal lines indicate 95% confidence intervals; numbers in parentheses indicate strong-positive patients. **Abbreviations:** ALSFRS-R, ALS Functional Rating Scale-Revised; Aβ, amyloid-beta; αSyn SAA, alpha-synuclein seed amplification assay; CC, complete case; ΔFS, disease progression rate from symptom onset to baseline; MI, multiple imputation; NfL, neurofilament light; %FVC, percent predicted forced vital capacity.

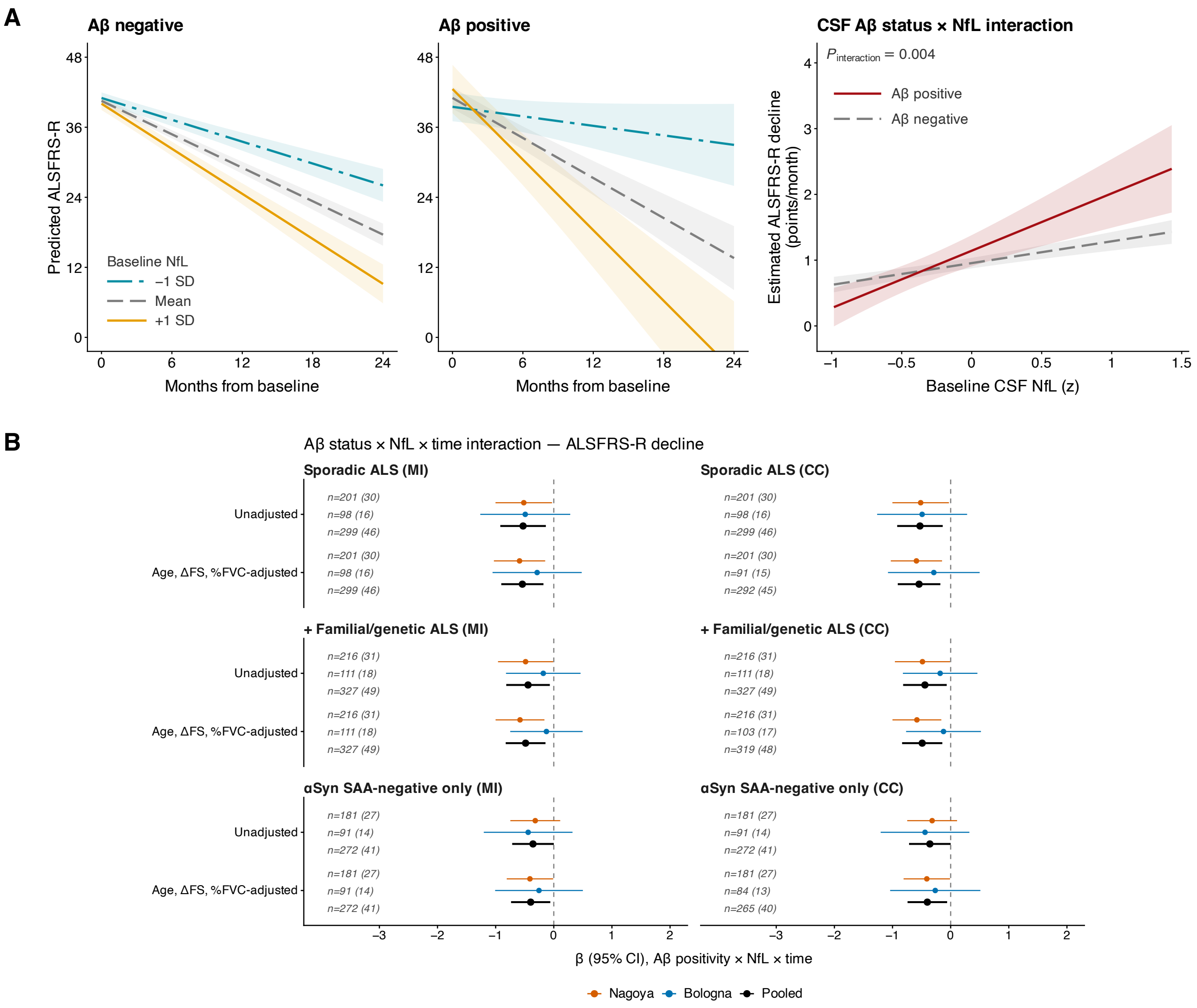


**Supplementary Fig. S5. Sensitivity analyses of LME models for Aβ status and ALSFRS-R decline**

**(A)** Predicted ALSFRS-R trajectories by CSF Aβ status at baseline CSF NfL of −1 SD, mean, and +1 SD (within-cohort z-scores; left and middle), and estimated monthly ALSFRS-R decline as a continuous function of baseline NfL (right), from pooled models in sporadic ALS adjusted for age, ΔFS, and %FVC (MI). **(B)** Aβ × NfL × time interaction coefficients (points/month per SD of NfL) for each cohort and the pooled cohort, in sporadic ALS (primary analysis), all ALS including familial and genetic cases, and sporadic ALS restricted to αSyn SAA negative subgroups, each with MI (left) and CC (right), unadjusted and adjusted for age, ΔFS, and %FVC. Lines and shaded bands indicate 95% confidence intervals. **Abbreviations:** ALSFRS-R, ALS Functional Rating Scale-Revised; Aβ, amyloid-beta; αSyn SAA, alpha-synuclein seed amplification assay; CC, complete case; CSF, cerebrospinal fluid; ΔFS, disease progression rate from symptom onset to baseline; MI, multiple imputation; NfL, neurofilament light; SD, standard deviation; %FVC, percent predicted forced vital capacity.
